# Oral Contraceptive Use Is Associated with Significant Differences in MicroRNA Cargo of L1CAM-Associated Extracellular Vesicles

**DOI:** 10.1101/2025.01.15.25320605

**Authors:** Dana M. Lapato, Rebekah Frye, Vasily Yakovlev, Roxann Roberson-Nay

## Abstract

Oral contraceptives (OCs) are approved for use after onset of menarche, which is well before brain maturation is complete. OC use may induce biochemical changes in the brain, especially during the neurobiologically dynamic adolescent/young adult years. MicroRNA cargo in L1CAM-associated extracellular vesicles was measured from serum samples collected from young women using the miRCURY LNA miRNA Focus PCR Panel (Qiagen) and validated using quantitative PCR. Linear regression and F-tests were applied to identify differentially expressed microRNAs by OC use (never versus current), and PANTHER pathway analysis was conducted on the gene targets of significantly differentially expressed microRNAs. Twelve microRNAs had significant differential expression variability by OC use (Bonferroni adjusted p < 0.002). Pathway analysis revealed that the 1254 unique genes targeted by the significant microRNAs were most enriched for the Gonadotropin-releasing hormone receptor pathway (FDR q = 5 × 10^−7^), which is associated with the release of gonadotropins, pubertal development, and reproduction. The results are consistent with the hypothesis that microRNA cargo in circulating extracellular vesicles may reflect brain-related biological activity and that OC use may influence extracellular vesicle cargo composition. The significant difference in expression variability may have implications for designing future studies, including power calculations.

## Introduction

Oral contraceptives (OCs) effectively treat many reproductive and hormone-related conditions, including endometriosis, abnormal or irregular uterine bleeding (e.g., menorrhagia), polycystic ovary syndrome, and migraines. Cross-sectional studies of adult women suggest that OC use is associated with structural and functional alterations to the brain^1–9^, but the mechanisms and directionality underlying these correlations remain uncertain. Consequently, significant knowledge gaps exist, including whether these changes are enduring and/or affect brain development. This paucity of research is surprising given that millions of young people are prescribed OCs every year.

Directly assaying cellular activity in the brain is not feasible, but monitoring changes in circulating extracellular vesicle (EV) cargo could offer a noninvasive approach to proxy biological states in inaccessible tissues like the brain. EVs are membrane-bound sacs that cross the blood-brain barrier and transport biologically active materials throughout the body to promote homeostasis and facilitate intercellular signaling. EVs are released by most cells, including brain cells, as a normal part of physiology^10–13^. Both the raw count of circulating EVs and EV cargo composition have been associated with pathophysiological features in traumatic brain injury, cancer, and neurodegenerative disorders like Alzheimer’s disease and Parkinson’s^11,14–18^. Relatively little is known about EVs during adolescence and early adulthood, but it is reasonable to hypothesize that they may be involved with signaling at least some of the systemic physiological changes during puberty. Moreover, given the potent impact estrogen can exert on gene expression and DNA methylation patterning, it is possible that OC use affects EV cargo composition, especially the microRNA component.

MicroRNAs (miRNAs) are short functional RNAs that regulate gene expression at the translational level by repressing messenger RNA (mRNA). A single miRNA can target multiple messenger RNAs (mRNAs), and individual mRNAs may be targeted by multiple miRNAs^19^. MiRNAs are abundant in EV cargo^20^ and suggest a plausible mechanism for long distance cell-to-cell feedback loops (i.e., Cell 1 sends miRNAs in an EV to Cell 2 to impact Cell 2’s gene regulation at the translational level^21^).

The purpose of this study was to test for differences in EV miRNA cargo during a narrow age range in early adulthood based on OC use. To improve the likelihood of identifying a brain-related biosignature, our analysis focused on L1CAM-associated exosomes. While not exclusively indicative of brain origin, L1CAM has been a useful cell-surface marker for brain-related research. Exosomes are an EV subgroup marked by small size (average diameter of ∼100 nm) and a collage of extracellular markers, including TSG101, CD63, CD81, and CD9^22^. EVs can carry a heterogeneous mixture of protein, DNA, and RNA cargo that putatively reflects the parent cell that secreted them^23^. They are taken up by distant cells, where they can affect cell function and behavior. Intercellular communication through exosomes seems to be involved in the pathogenesis of various disorders, including cancer, neurodegeneration, and inflammatory diseases. L1CAM-associated EV and exosome cargo have been consistently associated with neurodegenerative disorder status. L1CAM is a transmembrane cell-adhesion molecule highly expressed in neurons and a member of the immunoglobulin superfamily. While not exclusively expressed in neurons, L1CAM-associated EVs have been used extensively to identify and extract neuron-enriched EV subpopulations. Cargo contained in L1CAM-associated EVs may capture important insights in signaling pathways relevant to the developing brain, which has not yet been thoroughly explored.

## Methods

### Participants

Female participants were recruited between January 2022 and November 2022 in Richmond, Virginia. All participants were required to be at least 18 years old and generally healthy to be eligible for enrollment (see Supplemental File S1 for full exclusion criteria). Additionally, participants qualifying as OC controls could not currently be or ever have been prescribed hormone-based OC for any reason. Participants in the OC user group (hereafter referred to as “cases”) brought either their OC medication packet or pictures of the packet with them to the study visit so that study personnel could record the prescription name, progestin generation, number of days of active versus inactive pills, and synthetic hormone dosage (e.g., Loestrin® 24 FE, 1-24 active pill days, 25-28 inactive pill days, EE=0.02 mg/NEA=1.0 mg). After providing informed consent, all participants completed questionnaires and had a peripheral blood specimen collected by trained research staff. The study protocol was reviewed and approved by the VCU institutional review board (HM20020127).

### Exosome Extraction

Detailed protocols for exosome isolation and extraction can be found in the supplemental methods. Briefly, exosomes were extracted from serum samples by size exclusion chromatography method (35nm qEV Column, Izon) and precipitated overnight on a rotating mixer with L1CAM/CD171 Monoclonal Biotin Conjugated Antibody (eBio5G3 (5G3), Thermofisher). After overnight incubation samples were additionally incubated with Pierce™ Streptavidin Plus UltraLink™ Resin (Thermofisher). After several washings, precipitated L1CAM-associated exosomes were eluted. Our previous work demonstrated the high efficiency of this method for the extraction of the pure fraction of the intact L1CAM-associated exosomes, which was confirmed by Western blot analysis and transmission electron microscopy (TEM).^24^

### Exosomal RNA Isolation and RT-PCR

Total RNA (including miRNA) was isolated from exosome samples following the manufacturer’s instructions with the miRNeasy MicroRNA Extraction Kit (Qiagen), and then an equal amount of RNA from each sample was converted to cDNA using the miRCURY LNA RT Kit (Qiagen). The concentration and purity of the exosomal RNA samples were estimated using NanoDrop ND-1000 spectrophotometer (Thermo Scientific, Wilmington, DE). To improve power and increase the odds of finding generalizable signals, all cDNA samples were pooled into two groups corresponding to OC user and OC naive subgroups. MiRNAs were assayed using the Serum/Plasma Focus PCR panel (Qiagen), which is a commercially available microarray platform containing 179 LNA miRNA primer sets of miRNAs that have been commonly found in human plasma and serum. The amplification was performed in a QuantStudio 5 Real-Time PCR System (Thermo Fisher Scientific, USA), and the amplification curves were analyzed using the QuantStudio™ Design & Analysis Software v1.4.2, both for determination of count (Ct) values and for melting curve analysis. All assays were inspected for distinct melting curves, and temperatures were monitored and kept within the specifications for each assay. The 2-ΔΔCt method was used to calculate relative miRNA expression levels. MiRNAs that exhibited at least a 5-fold difference in normalized expression between OC case and control groups were subsequently assayed in all individual participants using quantitative PCR (qPCR).

### Quantitative Real Time PCR (qPCR) Data Analysis

Data from the qPCR experiment was imported into the R statistical environment^25^. Potential reference genes were identified by comparing the overall stability and relative expression of candidate miRNAs across OC cases and controls. Relative expression was calculated for each individual’s miRNAs by subtracting the reference gene expression value from all of their other miRNA counts and exponentiating that value (2^x). Group-based differences in means and variances were evaluated using the Wilcox test and F test, respectively, for OC users and OC naive controls based on normalized miRNA expression values. P values were corrected for multiple tests using Bonferroni adjustment (adjusted p = 0.05/26 tests = 0.002).

### Gene Set Enrichment Analysis

Significant miRNA gene targets were identified using mirTarBase^26^ and the resulting list of gene names was deduplicated and submitted to PANTHER (Protein ANalysis THrough Evolutionary Relationships; version 19.0)^27,28^ for overrepresentation analysis for biological processes, molecular functions, cellular components, and gene pathways. P-values were adjusted using False Discovery Rate (FDR)^29^.

## Results

Participant demographic characteristics by OC use are provided in Table 1. Hormone data validated self -reported OC use, with estrogen levels in OC users typically under 10 pg/mL (Table 1). All OC users were taking a combination pill that included ethinyl estradiol (range: 10-30 mcg) and various progestin components (i.e., Norethindrone Acetate=5, Desogestrel=2, Drospirenone=2, and Norgestrel=1). Progestin doses ranged 0.15 - 3.0 mg. Most OC users (85.7%) reported long term pill use (greater than 3 years).

**Table 1.** Demographic Table.

| Variable | OC Cases | OC Controls | p-value<sup>+</sup> |
| --- | --- | --- | --- |
| N | 10 | 10 | - |
| Age<sup>*</sup> | 19.01 (1.13) | 18.65 (1.07) | > 0.9 |
| *Self-reported race* |  |  |  |
| Asian | 1 | 1 |  |
| Black | 0 | 1 |  |
| Latina | 1 | 0 |  |
| White | 5 | 5 |  |
| *Hormones<sup>\*</sup>* |  |  |  |
| FSH | 2.81 (2.0) | 3.73 (1.9) | 0.289 |
| **LH** | **1.40 (1.6)** | **6.10 (6.2)** | **0.009** |
| Progesterone | 0.35 (0.3) | 1.56 (2.6) | 0.641 |
| **Estradiol** | **11.1 (1.6)** | **61.6 (39.9)** | **0.001** |

\* = Mean (SD); + Wilcoxon rank sum test
FSH = Follicle stimulating hormone; LH = Luteinizing hormone

Fourteen of the twenty participants with hormone measures had usable individual-level miRNA qPCR measures. Of the 179 miRNA probes assayed, 27 miRNAs were identified as candidates for individual-level measurement via qPCR. The overall gene expression patterns in OC users versus controls primarily differed in two ways. First, six miRNAs were expressed in OC users but largely absent from OC controls (miR-199a-3p, miR-148b-3p, miR-191-5p, miR-324-3p, miR-197-3p, miR-200c-3p). Second, miRNAs that were expressed in both OC users and controls tended to have less expression variability in OC users. Of those 27 miRNAs, one (miR-133a-3p) was selected to serve as a normalization control. Twelve of the remaining 26 (46%) miRNAs had statistically significant differences in gene expression variability (Figure 1). Given that variance-based tests are sensitive to outliers, the F tests were rerun only on expression values within 3 standard deviations of the mean expression value for each miRNA to identify the most robust variance differences. Seven of the 12 miRNAs remained significant in the sensitivity analysis. In total, the 12 statistically significant miRNAs target 1,254 unique genes according to annotations from MiRTarBase. N=114 of the genes (∼9.1%) were targeted by two or more of the significant miRNAs. None of the miRNAs exhibited a statistically significant difference in mean normalized expression after Bonferroni adjustment, although two miRNAs (miR-125b-5p, miR-126-5p) approached significance (adjusted p = 0.06 and 0.10, respectively). The n=1254 unique gene targets of miRNAs with significantly different variances by OC use were identified using miRTarBase. Gene set enrichment comparing these gene targets to all expressed human genes cataloged in the PANTHER database at the time of analysis (n=20580) identified 22 significantly enriched pathways (FDR q < 0.05). The most significantly enriched pathway was the Gonadotropin-releasing hormone receptor pathway (FDR q = 5 × 10^−7^; P06664), followed by the TGF-beta signaling pathway (FDR q = 1.3 × 10^−6^; P00052), the CCKR signaling map (FDR q = 6.0 × 10^−6^; P06959), and p53 pathway feedback loops 2 (FDR q = 6.4 × 10^−6^; P04398).

**Figure 1.**
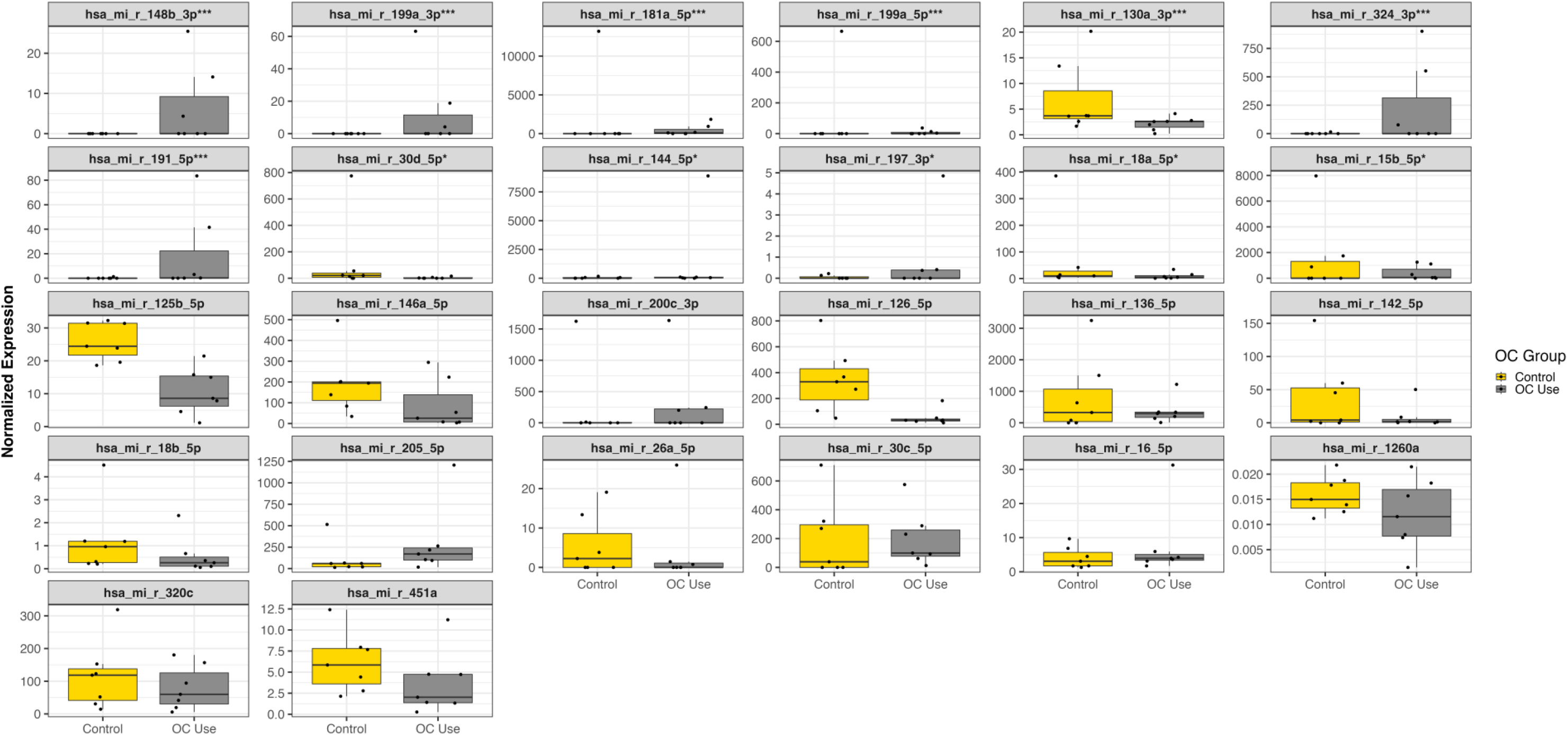
Normalized expression values were plotted by OC use for all microRNAs measured via quantitative PCR. Three asterisks (***) in the subplot heading indicates that the between-group difference in variance was statistically significant after Bonferroni adjustment and in the sensitivity analysis. A single asterisk (*) indicates that the result was significant in the initial analysis but not the sensitivity analysis. Note that each subplot has its own y-axis scale. A zero value for normalized expression means that the microRNA in question was not detected after 50 cycles of qPCR.

## Conclusions

There is still much to learn about the nature of EV microRNA cargo during development. Little is known about the normal expression patterns of extracellular vesicles or their cargo composition across the life course (or even simply across the menstrual cycle), how much variability exists within healthy populations, or what factors impact their expression patterns (e.g., sex [i.e., biological], exposure to air pollution [i.e., environmental], tobacco smoking [i.e., behavioral]). This study evaluated the impact of oral contraceptive use on EV microRNA expression in fourteen healthy women. Profound differences in expression variability were observed by oral contraceptive use in twelve miRNAs, and these small RNAs collectively target genes significantly enriched for twenty-two pathways after FDR correction. The top pathway result was the Gonadotropin-releasing hormone receptor pathway, which is intimately associated with the expression and regulation of gonadotropins (i.e., LH and FSH), pubertal development, and reproduction. This finding is consistent with OCs’ intended pharmacological action of inhibiting gonadotropin secretion. These results collectively support the notion that EV cargo may be valuable for monitoring development and may provide insight into sex-specific differences in development and post-pubertal risk liability for physical and mental health disorders. If reliable EV cargo proxies are identified, they could be useful biomarkers for monitoring the effects of OCs on the body (e.g., hypothalamic-pituitary-gonadal axis) and aiding in clinical assessments (e.g., OC efficacy and safety). Further, the observation of a subset of microRNAs expressed only in OC users raises the possibility that the use of synthetic hormones may evoke a unique gene expression profile. Previous studies have suggested that alcohol consumption can impact extracellular vesicle cargo. While alcohol use was assessed in this study, there was not enough statistical power to disentangle its potential impact in relation to OC use, so we will have to rely on future work to investigate possible interaction effects between OC use and behavioral traits like substance use.

Care must be taken when interpreting these results. On one hand, several factors speak to the credibility of the results. Though the sample size is modest, the analyses focus on a potent biochemical exposure and a narrow age range, which both likely bolster statistical power. Additionally, the extracellular vesicle samples were enriched for exosomes using immunoprecipitation, which may have further clarified the biological signal by reducing EV heterogeneity before microRNA measurement. The significant differences in microRNA expression variance were sizable in magnitude, survived Bonferroni correction, and most were robust to a conservative sensitivity analysis. Moreover, these microRNAs map on gene targets enriched for salient biological pathways, including the Gonadotropin-releasing hormone receptor pathway which is directly linked to the exposure of interest (i.e., oral contraceptives). At the same time, these results come from a modestly sized cohort with inherently limited diversity due to the small sample size. The individual-level microRNA results, in particular, should not be overinterpreted or considered credible until they are replicated. This caution is analogous to what would be exercised with genome-wide association study (GWAS) results, where the credibility of any individual result is not as robust as aggregate-level results (e.g., polygenic scores, pathway analyses), and replication is standard. Ideally, future studies will be able to enroll not only larger cohorts but also to apply sequencing technologies so that both known and novel microRNAs can be identified and quantified. An additional benefit of sequencing technologies is the potential to estimate genetic ancestry, dimensions of which could then be included as covariates in statistical models as is standard practice in GWAS. This step will be especially important if evidence emerges that extracellular vesicle cargo is influenced by genetic variation.

In conclusion, when planning future studies of extracellular vesicle cargo, researchers should take note of the potential for sizable differences in expression variability, which may be due not only to the trait or disorder of interest but also biological, environmental, or behavioral factors.

## Required Statements

### Data availability statement

All phenotypic measures, transcriptomic data, and annotation documents used or generated as part of this project have been deposited on the Open Science Framework (https://osf.io/pqdnx) under the Creative Commons Attribution-Noncommercial-ShareAlike 4.0 International license (CC BY-NC-SA 4.0). Supplemental details for the methods, including EV extraction and the pathway curation description for the Gonadotropin-releasing hormone receptor pathway from PANTHER, are available in Supplemental File 1.

### Ethics approval statement

This study was conducted in accordance with the ethical standards of the Declaration of Helsinki and was approved by the Institutional Review Board (IRB) at Virginia Commonwealth University. The protocol number for this study is HM20020127. All participants provided written informed consent prior to their inclusion in the study.

## Supplemental File S1 - Methods

### Comprehensive participant exclusion criteria

Participants were not eligible to enroll in this study if they endorsed any of the following conditions: 1) seizure disorder, 2) episode of psychosis, 3) serious, unstable medical condition, 4) inadequate production of human growth hormone, 5) congenital adrenal hyperplasia, 6) adrenal insufficiency, 7) lifetime pregnancy, 8) sex chromosome abnormality, 9) current weight that could likely impact endocrine functioning (BMI < 18 or BMI > 30), 10) current substance abuse/dependence, 11) history of traumatic brain injury, 12) lifetime cancer, and 13) contra-indications to MRI (e.g., implanted metal), and 24) any disorder not listed that impacts brain biology or endocrine system functioning.

### Expanded EV Methods

#### Extracellular Vesicle Isolation and Enrichment

Exosomes were extracted from serum samples by size exclusion chromatography method using 35nm qEVoriginal columns (Izon Science). Every 0.5 ml of serum yielded 1.5 ml of purified exosomes in 1xPBS (with no Mg) buffer. Exosome precipitation was performed overnight at +4°C on a rotating mixer with 8 μL of L1CAM/CD171 Monoclonal Biotin Conjugated Antibody (eBio5G3 (5G3), Thermofisher) in the presence of 3% BSA and 3x Proteinase/Phosphatase Inhibitors cocktail (Sigma-Aldrich) per 1.5 mL of purified exosomes in 1xPBS. After overnight incubation samples were additionally incubated with 40 μL Pierce™ Streptavidin Plus UltraLink™ Resin (Thermofisher) for 1 hour at +4°C. As a washing step, sample tubes were centrifuged with 500xg for 1 min, the supernatant was removed and saved as non-neuronal fraction of serum EVs, and resin beads were washed 2x times with 1 mL of pre-filtered (with 0.22μM filter) 1xPBS buffer. After the final wash, precipitated neuronal-enriched exosomes were lysed with 700 μL TRIzol Reagent (Invitrogen), total RNA was extracted using 200 μL chloroform, and precipitated via 2.5X volume of cold (−20C) ethanol with 5 μL of 20 mg/mL glycogen (ThermoFisher Scientific). RNA was then processed via the miRNeasy Serum/Plasma Advanced Kit microRNA Isolation Kit (Qiagen) using the manufacturer’s recommendations.

#### Exosomal RNA Isolation and RT-PCR

Total RNA was isolated from the exosomal samples following the manufacturer’s instructions with the miRNeasy MicroRNA Extraction Kit (Qiagen) and then converted to cDNA using the miRCURY LNA RT Kit (Qiagen). The samples with total exosomal RNA and cDNA were stored in a freezer at −80°C. The concentration and purity of the exosomal RNA samples were estimated using NanoDrop ND-1000 spectrophotometer (Thermo Scientific, Wilmington, DE).

All cDNA samples were pooled into two groups corresponding to OC users and OC naive controls and then assayed using the Serum/Plasma Focus PCR panel (Qiagen). This commercially available microarray platform contains 179 LNA miRNA primer sets of miRNAs that have been commonly found in human plasma. The Serum/Plasma Focus miRNA PCR Panels include potential reference genes and probes for evaluating potential sample contamination or damage (e.g., hemolysis). Each PCR panel also contains set of the negative control (H_2_O), five sets of RNA Spike-In controls to evaluate RNA extraction (Spike-Ins 2-4-5; concentration ratio 1:100:10000), and cDNA synthesis control (Spike-In 6) and an internal control to perform inter-plate calibration (Spike-In 3). The amplification was performed in a QuantStudio 5 Real-Time PCR System (Thermo Fisher Scientific, USA). Samples were amplified using RT2 SYBR®Green qPCR Mastermix with ROX (carboxy-X-rhodamine) passive reference dye from QIAGEN. The real-time PCR data were normalized by ROX passive reference. The amplification curves were analyzed using the QuantStudio™ Design & Analysis Software v1.4.2, both for determination of count (Ct) values and for melting curve analysis. All assays were inspected for distinct melting curves and the melting temperature was checked to be within known specifications for each assay. The 2-ΔΔCt method was used to calculate relative miRNA expression levels.

### Background on the Gonadotropin-releasing hormone receptor pathway from PANTHER

Information about the methods and resources used to annotate and describe the Gonadotrophin-releasing hormone receptor pathway (accession ID: P06664) is provided on the PANTHER website and copied below for reference.

#### PANTHER pathway accession ID: P06664

The GnRH receptor (GnRHR), expressed at the cell surface of the anterior pituitary gonadotrope is critical for normal secretion of gonadotropins LH and FSH, pubertal development, and reproduction. The signaling network downstream of the GnRHR and the molecular bases of the regulation of gonadotropin expression have been the subject of intense research. The murine LbetaT2 cell line represents a mature gonadotrope, and therefore is an important model for the study of GnRHR signaling pathways, and modulation of the gonadotrope cell by physiological regulators. In order to facilitate access to the information contained in this complex and evolving literature, we have developed and curated a comprehensive knowledgebase of the GnRHR signaling in the LbetaT2 cell in the form of a process diagram (Fink et al., 2010; PMID: 20592162). Positive and negative controls of gonadotropin gene expression, which included GnRH itself, hypothalamic factors, gonadal steroids and peptides, as well as other hormones, were illustrated. The GnRHR signaling pathway is being updated yearly, based on the latest publications in the field in addition to experts’ suggestions. The pathway map was curated using CellDesigner ver.4.1 (http://celldesigner.org/). For simplification purposes, activating reactions, including those linking transcription factors to genes, were depicted with “State Transition” black filled arrows. Inhibitory reactions were colored in red. The size and color of each module were configured by us. Briefly, round angle green squares signify cell signaling proteins, whereas green and yellow circles represent small molecules and ions, respectively; round angle blue squares symbolize transcription factors, and round angle purple squares strictly correspond to nuclear receptors; yellow/green rectangles designate genes, on which some response elements were represented as small white squares. The following cellular compartments were illustrated on the diagram, as indicated: cytoplasm, endoplasmic reticulum, proteasome, and nucleus. Please send your feedback/comments/suggestions to the GnRHR knowledgebase curator Hanna Pincas.

